# Evaluating community pharmacists’ involvement in patient counselling and health education in Nairobi, Kenya: A cross-sectional study

**DOI:** 10.1101/2025.03.31.25324939

**Authors:** Manjari Seraphine Wanjiro, Ermias Mergia Terefe

## Abstract

**Background:** Community pharmacists serve as the most accessible healthcare providers, playing a pivotal role in patient counselling, health education, and disease prevention. Despite their critical function in the healthcare system, limited research has examined their actual involvement in these areas within Nairobi, Kenya. This study aims to assess community pharmacists’ engagement in patient counselling and health education, comparing self-reported practices with observed behaviours using a simulated patient approach.

**Methods:** A cross-sectional study was conducted among registered community pharmacies in Nairobi. A total of 60 community pharmacies were randomly selected, with 48 community pharmacists completing the study. Data collection involved two methods: structured questionnaires administered to pharmacists assessing self-reported patient counselling practices and simulated patient visits evaluating actual pharmacist-patient interactions. Three case scenarios (diabetes medication request without prescription, over-the-counter acne treatment request, and severe acne consultation) were used to assess pharmacists’ counselling competence, adherence to regulations, and patient education efforts. Descriptive statistics were applied to analyse the data, comparing self-reported responses with observed practices.

**Results:** Findings revealed significant discrepancies between pharmacists’ self-reported counselling practices and their actual interactions with patients. While, 66.67% of pharmacists claimed to provide detailed medication counselling, yet only 26.67% of simulated patients received adequate guidance. Additionally, 75% of pharmacists provided minimal verbal counselling beyond stating the drug name and dosage. Patient satisfaction levels were low, with most simulated patients rating their encounters poorly on a 20-point scale.

**Conclusion:** The study underscores critical gaps in patient counselling, health education, and adherence to prescription regulations among community pharmacists in Nairobi. The disconnect between self-reported and observed counselling practices suggests the need for targeted training programs to enhance pharmacists’ communication skills, regulatory compliance, and disease prevention counselling. Strengthening enforcement mechanisms and integrating patient-centred education strategies in pharmacy curricula are essential to optimizing pharmacists’ roles in healthcare. Future research should explore barriers to effective counselling and evaluate the impact of training interventions on improving pharmacists’ patient engagement and adherence to professional standards.

## 1. Introduction

Community pharmacists (CPs) serve as the first point of contact for many patients, offering easily accessible healthcare services without the need for appointments or referrals(1),(2). Their geographical distribution and relatively lower costs compared to hospital consultations make them an integral part of the healthcare system, particularly in low-resource settings. During dispensing medications, CPs are responsible for ensuring the rational and effective use of drugs by counselling patients on medication adherence, potential side effects, and safety precautions(3). However, despite their strategic role in patient care, studies indicate that CPs often fail to provide adequate counselling, focusing instead on the transactional aspects of medication dispensing (4),(5).

Health education and promotion are essential components of public health, enabling individuals to take charge of their well-being by fostering health-conscious behaviours (6). Community pharmacists are uniquely positioned to engage in health promotion initiatives, particularly for chronic conditions such as diabetes and hypertension, which are on the rise due to poor lifestyle choices(7). Through structured educational programs, CPs can provide valuable guidance on medication adherence, lifestyle modifications, and disease prevention, ultimately improving population health outcomes(7). However, the extent to which CPs are actively involved in these activities remains unclear, particularly in Kenya, where data on their contributions to health education and promotion is scarce.

Traditionally, pharmacists have been viewed primarily as dispensers of medications. The evolving healthcare landscape necessitates a shift toward patient-centred care, where pharmacists actively engage in providing guidance on medication use, disease prevention, and lifestyle modifications (1).Inadequate counselling skills among CPs lead to suboptimal treatment outcomes and medication-related problems(1). Studies conducted in different settings have demonstrated that pharmacists often fail to adequately counsel patients, resulting in poor medication adherence and ineffective disease management. Despite existing guidelines emphasizing the importance of patient counselling, there remains a gap between the expected and actual practice among CPs.

Several studies have highlighted that, although pharmacists recognize the importance of patient education, their actual involvement in counselling and health promotion is limited. The reasons for this include lack of time, inadequate training, and minimal enforcement of counselling regulations (8). Inadequate pharmacist-patient interaction can contribute to medication misuse, therapeutic failures, and avoidable adverse effects. Furthermore, in developing countries such as Kenya, limited research has been conducted to assess CPs’ engagement in health promotion and disease prevention efforts. Understanding these gaps is essential for informing policy decisions and designing training programs to enhance pharmacists’ roles in public health.

This study aims to assess the level of involvement of community pharmacists in patient counselling, health education, and health promotion in Nairobi, Kenya. Specifically, the study seeks to evaluate patient counselling practices, determine CPs’ engagement in health promotion initiatives, assess the quality of patient education provided, and identify barriers hindering effective counselling and health education. The findings will contribute to bridging the existing gaps in pharmaceutical practice and provide evidence to support regulatory enforcement and policy development aimed at optimizing pharmacists’ roles in healthcare.

## 2. Methods

### Study Design

This study utilized a qualitative descriptive research design to evaluate the extent of community pharmacists’ (CPs) involvement in patient counselling, health education, and health promotion(9). The study sought to compare self-reported practices of CPs with actual pharmacist-patient interactions observed through simulated patient visits. Across-sectional survey was conducted in parallel with the simulated patient method to provide a comprehensive assessment of pharmacists’ engagement in these key aspects of healthcare delivery (10).

### Study Setting

The study was conducted across all 17 constituencies in Nairobi, Kenya, between November 2022 and February 2023. Community pharmacies included in the study were selected based on their registration status and active licensure with the Pharmacy and Poisons Board (PPB). The study location was chosen due to the high density of pharmacies and the significant role they play in primary healthcare within urban settings.

### Study Population and Sampling

#### Target Population

The study population comprised all community pharmacies registered with the PPB and holding active operational licenses in Nairobi County as of September 2022. According to PPB records, there were 1,361 registered pharmacies in Nairobi, of which 1,213 met the study’s inclusion criteria.

### Eligibility Criteria

#### Inclusion Criteria

- Community pharmacies officially registered with the PPB in Nairobi.
- Pharmacies with valid and active operational licenses at the time of data collection

#### Exclusion Criteria

- Pharmacies that were inactive or not in operation during the study period
- Community pharmacists who declined participation

### Sampling Design

#### Sampling Frame

The sampling frame consisted of 1,213 registered and operational community pharmacies in Nairobi County. This frame was used to systematically select the study sample, ensuring that the findings would be representative of the broader population of community pharmacies within the county.

#### Sampling Technique

A stratified random sampling approach was employed to ensure equitable representation across all 17 constituencies in Nairobi. Pharmacies were first categorized into clusters based on their respective constituencies. A proportional random sampling strategy was then applied to select a representative number of pharmacies from each cluster. This method facilitated an unbiased selection process, thereby enhancing the reliability and generalizability of the study findings (9).

#### Sample Size Determination

The sample size was determined using a statistical approach appropriate for large populations (11). Given that the total population of registered community pharmacies in Nairobi stood at 1,213, a representative sample was calculated to maximize precision and reliability. Adjustments for population size were applied, resulting in a final sample of 60 pharmacies. This sample size was deemed sufficient to capture variations in pharmacists’ patient counselling and health education practices while ensuring the generalizability of the results.

### Data Collection

#### Survey Instrumentation

Data collection was conducted using structured questionnaires and scorecards. The questionnaires were administered to community pharmacists to evaluate their involvement in patient counselling, health education, and health promotion. In parallel, scorecards were provided to simulated patients, who assessed the quality of pharmacist-patient interactions based on predefined criteria.

#### Pilot Study

Prior to the main study, a pilot study was carried out to evaluate the feasibility and clarity of the research instruments, including the questionnaires and selected case scenarios. Feedback from the pilot phase led to refinements, such as eliminating ambiguous or repetitive questions, ensuring clarity in wording, and improving overall study design (12).

#### Cross-Sectional Survey

A structured questionnaire was distributed to the selected community pharmacists. The questionnaire consisted of three main sections: demographic information, patient counselling practices, and pharmacists’ involvement in health education and promotion. Responses were measured using a three-point Likert scale (always discussed, discussed sometimes, and never discussed) to assess the extent of pharmacist engagement in these key areas.

#### Simulated Patient Method

To complement the cross-sectional survey, a simulated patient approach was utilized. Three senior pharmacy students (5th-year pharmacy students) were recruited and trained to portray specific case scenarios requiring pharmacist intervention. The case scenarios focused on diabetes mellitus and acne vulgaris, as these conditions necessitate thorough patient counselling, health education, and wellness promotion alongside pharmacological management.

A structured checklist was developed to systematically evaluate pharmacists’ interactions with simulated patients. The checklist assessed key elements such as patient history-taking, provision of appropriate prescriptions, medication counselling, and health promotion strategies (13). The parameters on the checklist mirrored those in the pharmacist-administered questionnaires, allowing for direct comparison between self-reported practices and observed behaviours.

### Case Scenarios

#### Scenario 1: Diabetes Medication Request Without Prescription

Simulated patient 1 visited community pharmacies requesting Treviamet (sitagliptin/metformin) 50/1000 mg without presenting a prescription or offering additional information unless prompted by the pharmacist. Upon further inquiry, the simulated patient provided the following details:

- The medication was intended for personal use.
- They had been diagnosed with diabetes 14 months prior.
- Their last HbA1C test, conducted seven months ago, was 13%.
- A recent fasting blood glucose test (three weeks ago) showed a reading of 9 mmol/L.
- Their last medical review was seven months ago.

Pharmacists were assessed based on their ability to identify the need for a prescription, provide proper counselling on diabetes management, and offer lifestyle modification advice.

#### Scenario 2: Over-the-Counter Acne Treatment Request

Simulated patient 2 visited community pharmacies requesting benzoyl peroxide and tretinoin for acne treatment. When questioned by the pharmacist, the patient provided the following information:

- The medication was for personal use.
- The patient had inflammatory acne.
- No previous treatments had been attempted.

Pharmacists were evaluated on their willingness to provide counselling on proper acne treatment, correct medication use, potential side effects, and lifestyle modifications to prevent acne flare-ups.

#### Scenario 3: Severe Acne Consultation

Simulated patient 3 presented at the community pharmacies seeking pharmacist guidance on managing severe inflammatory acne. The patient provided the following details upon further questioning:

- They had been experiencing recurrent acne for the past 24 months.
- Acne flare-ups lasted about two months before subsiding.
- They had previously used tretinoin and benzoyl peroxide but found them ineffective.
- They had only used medications during flare-ups, discontinuing treatment once improvements were noticed or when acne worsened during therapy.

Pharmacists were assessed based on their ability to take a thorough patient history, provide appropriate counselling on acne treatment adherence, and recommend either over-the-counter treatments or referral to a dermatologist if necessary.

Each simulated patient interaction was evaluated using a structured checklist assessing pharmacists’ inquiry about patient history, provision of counselling, medication guidance, and health education efforts.

### Ethical Considerations

This study adhered to ethical research principles, ensuring participant confidentiality and obtaining informed consent. The study received approval from the United States International University-Africa (USIU-A) Institutional Review Board (IRB) approval number USIU-A/IRB/F068 and the National Commission for Science, Technology, and Innovation (NACOSTI) licence number NACOSTI/P/22/22465. To maintain anonymity, the names and locations of pharmacies were not recorded, and simulated patients were identified numerically.

### Data Analysis

Collected data were coded and entered into Microsoft Excel, then analysed using SPSS version 18. Descriptive statistics such as frequencies and percentages were used to summarize findings. Data from self-reported surveys and simulated patient checklists were compared to identify discrepancies between reported and actual pharmacist practices.

## 3. Results

### Demographic Characteristics of Participants

A total of 60 community pharmacies were included in the study, with 48 community pharmacists responding to the survey. Among the respondents, 55%(n=26) held diplomas in pharmacy, while 45%(n=22) possessed a bachelor’s degree in pharmacy. The majority (68%) had over five years of experience in community pharmacy practice, whereas 32%(n=15) had been in practice for fewer than five years. The study also revealed that 72% (n=35) of pharmacists worked over 10 hours per day, reflecting the high workload and demanding nature of community pharmacy operations in Nairobi. These findings underscore the potential impact of long working hours on pharmacists’ ability to engage in patient counselling and health education effectively.

### Pharmacists’ Self-Reported Practices in Patient Counselling and Health Education

Survey responses indicated that 71% (n=34) of pharmacists reported routinely inquiring about patient history, including medication history, symptoms, and allergies, before dispensing medication. Additionally, 66.67% (n=32) of pharmacists claimed to provide comprehensive drug information, covering indications, dosage, contraindications, drug interactions, and potential side effects. However, only 50%(n=24) reported actively engaging in patient education on disease prevention and non-pharmacological treatment approaches, such as lifestyle modifications for chronic conditions like diabetes and hypertension. The findings suggest that while pharmacists acknowledge the importance of patient counselling, its implementation remains inconsistent.

### Findings from Simulated Patient Interactions

#### Scenario 1: Diabetes Medication Request Without Prescription

In the diabetes management scenario, where a simulated patient requested Treviamet (sitagliptin/metformin) without a prescription, only 40% (n=19) of pharmacists adhered to regulatory guidelines by not dispensing the medication without a valid prescription. The remaining 60% (n=29) either dispensed the medication without further inquiry or asked minimal follow-up questions before proceeding. Among those who engaged in further questioning, only 35% (n=17) inquired about prior diabetes management and adherence to prescribed therapy, while 25%(n=12) provided counselling on the role of lifestyle modifications such as diet and exercise in managing diabetes. These findings highlight a gap in adherence to dispensing regulations and the provision of patient-centred diabetes care.

#### Scenario 2: Over-the-Counter Acne Treatment Request

In the acne treatment scenario, 48% (n=23) of pharmacists correctly identified the appropriate first-line treatment and provided counselling on the use of benzoyl peroxide and tretinoin. However, 36% (n=17) recommended non-specific analgesics or alternative skincare products without clear justification, and 16% (n=8) failed to provide structured counselling on acne management. Notably, only 30% (n=14) of pharmacists discussed additional lifestyle modifications, such as proper skincare hygiene and dietary considerations for acne control. This lack of comprehensive counselling may contribute to suboptimal treatment outcomes for patients managing acne-related concerns.

#### Scenario 3: Severe Acne Consultation

For the simulated patient presenting with severe inflammatory acne, 55% (n=26) of pharmacists recommended continuing the use of over-the-counter treatments, despite the patient reporting poor results with previous medications. Only 20% (n=10) of pharmacists advised the patient to seek medical consultation with a dermatologist for further evaluation and treatment. Additionally, 45%(n=22) provided limited counselling, focusing solely on product selection without discussing adherence, potential side effects, or alternative treatment approaches. The findings suggest that many pharmacists may lack the confidence or knowledge to provide comprehensive counselling for complex dermatological conditions, underscoring the need for additional training in this area.

#### Comparison of Self-Reported Practices and Simulated Patient Observations

A significant discrepancy was observed between pharmacists’ self-reported practices and their actual counselling behaviour during simulated patient visits. While 71% (n=34) of pharmacists claimed to routinely inquire about patient history, only 15.26% (n=7) actively engaged in a thorough patient assessment when observed in practice. Similarly, although 66.67% (n=32) of pharmacists reported providing comprehensive drug information, only 26.67% (n=13) of simulated patients received adequate guidance regarding medication use. These inconsistencies highlight a gap between perceived and actual pharmacist performance in patient counselling and health education.

#### Patient Satisfaction and Medication Labelling

Simulated patients assessed their interactions with pharmacists based on communication clarity, counselling effectiveness, and overall satisfaction. The results showed that 59% (n=28) simulated patients rated their pharmacist encounters between 0-5 on a 20-point scale, indicating a generally low level of satisfaction. Additionally, 75% (n=36) of pharmacists failed to provide verbal counselling beyond stating the drug name and dosage, while 64% (n=31) of prescriptions lacked critical labelling details, including the patient’s name, dosage schedule, and administration instructions. The lack of comprehensive labelling and verbal counselling poses a potential risk to medication safety and adherence.

## 4. Discussion

The findings of this study highlight significant discrepancies between community pharmacists’ self-reported counselling practices and their actual observed behaviour during simulated patient visits. While pharmacists generally acknowledged the importance of patient counselling and health education, the study revealed notable inconsistencies in their implementation. These results suggest the need for targeted interventions to enhance pharmacists’ engagement in patient-centred care.

### Discrepancies Between Self-Reported and Observed Practices

One of the most striking findings was the gap between pharmacists’ self-reported practices and their actual counselling behaviour. Although 71% (n=34) of pharmacists claimed to routinely ask about patient history before dispensing medication, only 15.26% (n=7) were observed conducting thorough patient assessments. This discrepancy is concerning, as proper history-taking is fundamental to ensuring medication safety and efficacy. Similarly, while 66.67% (n=32) of pharmacists reported providing comprehensive drug information, only 26.67% (n=13) of simulated patients received adequate counselling. These findings align with previous studies that indicate that pharmacists often overestimate their engagement in patient education, possibly due to high workloads, time constraints, or lack of structured patient interaction protocols.

### Regulatory Adherence and Ethical Considerations in Medication Dispensing

A particularly concerning observation was the non-adherence to prescription regulations. In the diabetes management scenario, 60% (n=29) of pharmacists dispensed Treviamet without a valid prescription, despite being prescription only medicine. This practice raises significant ethical and regulatory concerns, as improper dispensing of antidiabetic medications can lead to adverse health outcomes due to inappropriate dosing, drug interactions, or patient non-adherence. The lack of rigorous questioning and documentation before dispensing prescription medications suggests that stronger enforcement of regulatory guidelines and pharmacist training on ethical dispensing practices are needed.

### Limitations in Disease Prevention and Non-Pharmacological Counselling

The study also highlighted gaps in pharmacists’ engagement in preventive health counselling and non-pharmacological management. While half of the pharmacists reported providing education on disease prevention, lifestyle modifications, and non-pharmacological interventions, only 30% of simulated patient encounters had such advice. This lack of engagement is particularly concerning in the management of chronic diseases such as diabetes and hypertension, where lifestyle modifications play a crucial role in treatment outcomes. Additionally, the limited discussion of lifestyle factors in acne management suggests a need for improved pharmacist training in dermatological conditions and patient education strategies. Although these findings show an improvement those found by Santos et al., (2013) where none of the pharmacists involved in the study educated simulated patients on drug interactions, adverse reaction and any non-pharmacological alternatives there is more that can be done.

### Pharmacists’ Role in Dermatological Counselling

The findings related to acne management further reinforce the need for enhanced pharmacist training. Although nearly half (48%) of pharmacists correctly identified appropriate first-line treatments for acne, a substantial proportion (36%) recommended non-specific analgesics or alternative skincare products without clear justification. Furthermore, only 20% of pharmacists recommended referral to a dermatologist for patients presenting with severe acne. This indicates a lack of confidence or knowledge regarding appropriate referral pathways and comprehensive dermatological counselling, underscoring the necessity of continued professional development in this area.

### Patient Satisfaction and Medication Labelling Deficiencies

Patient satisfaction scores revealed widespread dissatisfaction with pharmacist-patient interactions. A majority of simulated patients rated their encounters between 0-5 on a 20-point scale, indicating poor communication and counselling quality. One of the most alarming findings was that only 13% of patients received properly labelled medications. The absence of critical details such as dosage instructions, contraindications, and administration guidelines on prescription labels increases the risk of medication errors and poor adherence. Moreover, 75% of pharmacists failed to provide verbal counselling beyond stating the medication name and dosage, further contributing to suboptimal therapeutic outcomes.

### Implications for Policy and Practice

The findings of this study have significant implications for pharmacy practice, regulatory bodies, and pharmacy education. There is an urgent need for continued professional development programs to enhance pharmacists’ competency in patient counselling, particularly in chronic disease management, dermatological conditions, and lifestyle modification strategies. Regulatory agencies must also strengthen enforcement measures to ensure adherence to prescription guidelines and improve medication labelling standards. Additionally, pharmacy education curricula should place greater emphasis on communication skills and patient-centred care approaches to better prepare future pharmacists for their roles in healthcare delivery.

### Recommendations for Future Research

Further research is needed to explore the barriers preventing pharmacists from fully engaging in patient counselling and education. Qualitative studies could provide deeper insights into pharmacists’ perceptions of their roles, workload challenges, and potential strategies for improving counselling practices. Additionally, intervention studies evaluating the effectiveness of targeted training programs in enhancing pharmacists’ counselling skills and regulatory compliance would be valuable in guiding future practice improvements.

## 5. Conclusion

This study highlights the crucial role of community pharmacists in patient counselling, health education, and medication safety. While pharmacists serve as frontline healthcare providers, the findings reveal significant gaps between their self-reported practices and actual interactions with patients. Despite many pharmacists acknowledging the importance of counselling and education, simulated patient visits demonstrated inconsistencies in history-taking, medication counselling, and adherence to prescription regulations.

The lack of comprehensive patient assessments and inadequate provision of medication-related information raise concerns about patient safety and therapeutic outcomes. Additionally, deficiencies in medication labelling and verbal counselling further contribute to the risk of medication errors and poor adherence. These findings underscore the need for structured interventions aimed at improving pharmacists’ engagement in patient-centred care.

To address these gaps, targeted pharmacist training programs should be implemented to enhance communication skills, regulatory compliance, and disease prevention counselling. Strengthening enforcement mechanisms for prescription regulations is equally critical to ensure adherence to ethical dispensing practices. Furthermore, pharmacy education curricula should place greater emphasis on practical counselling techniques and real-world patient interactions.

Improving pharmacists’ involvement in health education and disease prevention is essential for optimizing healthcare delivery in Nairobi and beyond. Future research should explore the barriers that hinder pharmacists from fully engaging in patient counselling and evaluate the effectiveness of training interventions designed to bridge these gaps. By prioritizing pharmacist training, policy enforcement, and patient-centred education strategies, community pharmacies can significantly contribute to improved medication safety and public health outcomes.

## Data Availability

All relevant data are within the manuscript and its Supporting Information files.

## Acknowledgments

We would like to appreciate the willingness of supervisors, community pharmacists and simulated patients for their vital role in this study.

## Notes

### Competing Interest Statement

The authors have declared no competing interest.

### Funding Statement

The author(s) received no specific funding for this work.

### Author Declarations

United States International University-Africa (USIU-A) Institutional Review Board (IRB) approval number USIU-A/IRB/F068 and the National Commission for Science, Technology, and Innovation (NACOSTI) licence number NACOSTI/P/22/22465.

## REFERENCES.

1. Osemene KP, Ihekoronye RM, Jegede A. Assessing Counseling Practices of Community Pharmacists in Nigeria. Vol. 23, East and Central African Journal of Pharmaceutical Sciences. 2020.

2. Palaian S, Alomar M, Hassan N, Boura F. Opportunities for extended community pharmacy services in United Arab Emirates: perception, practice, perceived barriers and willingness among community pharmacists. J Pharm Policy Pract. 2022;15(1):1– 10.

3. Netere AK, Erku DA, Sendekie AK, Gebreyohannes EA, Muluneh NY, Belachew SA. Assessment of community pharmacy professionals’ knowledge and counseling skills achievement towards headache management: a cross-sectional and simulated-client based mixed study. J Headache Pain [Internet]. 2018 Dec 16;19(1):96. Available from: https://thejournalofheadacheandpain.biomedcentral.com/articles/10.1186/s10194-018-0930-7

4. Al Qarni H, Alrahbini T, Alqarni AM, Alqarni A. Community pharmacist counselling practices in the Bisha health directorate, Saudi Arabia-simulated patient visits. BMC Health Serv Res. 2020;20(1):1–7.

5. Showande SJ, Laniyan MW. Patient medication counselling in community pharmacy: evaluation of the quality and content. J Pharm Policy Pract [Internet]. 2022 Dec 16;15(1):103. Available from: https://joppp.biomedcentral.com/articles/10.1186/s40545-022-00502-3

6. Caron RM, Noel K, Reed RN, Sibel J, Smith HJ. Health Promotion, Health Protection, and Disease Prevention: Challenges and Opportunities in a Dynamic Landscape. AJPM Focus [Internet]. 2024 Feb;3(1):100167. Available from: https://linkinghub.elsevier.com/retrieve/pii/S2773065423001049

7. Okoro RN, Nduaguba SO. Community pharmacists on the frontline in the chronic disease management: The need for primary healthcare policy reforms in low and middle income countries. Explor Res Clin Soc Pharm [Internet]. 2021 Jun;2:100011. Available from: https://linkinghub.elsevier.com/retrieve/pii/S2667276621000111

8. Adepu R, Nagavi B. Attitudes and behaviors of practicing community pharmacists towards patient counselling. Indian J Pharm Sci [Internet]. 2009;71(3):285. Available from: http://www.ijpsonline.com/text.asp?2009/71/3/285/56029

9. Asenahabi BM. Basic of research design: a guide to selecting appropriate research design. Int J Contemp Appl Res [Internet]. 2019;6(5):76–89. Available from: www.ijcar.net

10. Surur AS, Getachew E, Teressa E, Hailemeskel B, Getaw NS, Erku DA. Self-reported and actual involvement of community pharmacists in patient counseling: a cross-sectional and simulated patient study in Gondar, Ethiopia. Pharm Pract (Granada) [Internet]. 2017 Mar 31;15(1):890–890. Available from: https://www.pharmacypractice.org/index.php/pp/article/view/890

11. Charan J, Biswas T. How to calculate sample size for different study designs in medical research? Indian J Psychol Med. 2013;35(2):121–6.

12. Alaqeel S, Abanmy NO. Counselling practices in community pharmacies in Riyadh, Saudi Arabia: a cross-sectional study. BMC Health Serv Res [Internet]. 2015 Jun 15;15(1):557. Available from: 10.1186/s12913-015-1220-6

13. Hamadouk RM, Arbab AH, Yousef BA. Assessment of Community Pharmacist’s Practice and Patient Counselling Toward Acute Diarrhea Treatment in Khartoum Locality: A Simulated Patient Study. Integr Pharm Res Pract [Internet]. 2021 Nov;Volume 10:145–52. Available from: https://www.dovepress.com/assessment-of-community-pharmacists-practice-and-patient-counselling-t-peer-reviewed-fulltext-article-IPRP

14. Santos AP, Mesquita AR, Oliveira KS, Lyra Jr DP. Assessment of community pharmacistś counselling skills on headache management by using the simulated patient approach: a pilot study. Pharm Pract. 2013;11(1):3–7.

